# The gSOS Polygenic Score is Associated with Bone Density and Fracture Risk in Childhood

**DOI:** 10.1101/2025.04.22.25325387

**Authors:** Jonathan A. Mitchell, Jonathan Bradfield, Shana E. McCormack, Alessandra Chesi, Heidi J. Kalkwarf, Joan M. Lappe, Sharon E. Oberfield, Dana L. Duren, John A. Shepherd, Kurt D. Hankenson, Andrea Kelly, Hakon Hakonarson, Struan F.A. Grant, Babette S. Zemel

## Abstract

The polygenic risk score *genetic quantitative ultrasound speed of sound (gSOS)* was developed using machine learning algorithms in adults of European ancestry and associates with reduced odds of fracture in adults. We aimed to determine if gSOS was associated with bone health in children.

Two observational studies of children were evaluated: (1) children enrolled in the Bone Mineral Density in Childhood Study (BMDCS) with genetic data (N=1,727); and (2) children with genetic data for research at the Children’s Hospital of Philadelphia (CHOP; N=10,301).

Genetic variants were used to calculate gSOS and genetic ancestry. For the BMDCS, puberty stage, dietary calcium, physical activity and fracture accumulation (none or ≥1 fracture) were self-reported, height and weight were measured and BMI calculated. Areal bone mineral density (aBMD) of the lumbar spine, hip, radius, and whole body were assessed by dual energy X-ray absorptiometry and expressed as Z-scores. The CHOP study paired genetic data with documentation of fracture in the electronic health record (EHR).

gSOS associated with higher aBMD Z-scores across 7 skeletal sites [e.g., a 1 SD increase in gSOS associated with 0.17 (95% CI: 0.10-0.24) higher lumbar spine aBMD Z-score]. These associations were consistent for males and females, age, puberty stage, and lifestyle factors, and most consistent among children of European genetic ancestry. A 1 SD increase in gSOS associated with 12% and 16% reduced likelihood of self-reported fracture in the BMDCS (OR=0.84, 95% CI: 0.74, 0.95) and a recorded fracture in the CHOP EHR (OR=0.88; 95% CI: 0.82, 0.95). No sex or genetic ancestry differences were found.

A higher gSOS score associated with higher aBMD at multiple skeletal sites and reduced odds of fracture in two independent pediatric samples. This genetic tool may have clinical utility to help enhance bone health in early life and protect against fracture across the lifespan.

**Lay summary:** In adults, the polygenic risk score gSOS associates with reduced fracture risk. This study evaluates the relationship of gSOS to bone density and fractures in two groups of children. We found that a 1 standard deviation increase in gSOS was associated with higher bone density at multiple skeletal sites. In our two groups of children, a 1 standard deviation increase in gSOS associated with reduced odds of fracture in children by 12% (95% CI: 0.82, 0.95) and 26% (95% CI: 0.74, 0.95). Having a higher gSOS may enhance bone accretion in early life, and protect against fracture across the lifespan.

## Introduction

Osteoporosis is a debilitating disease characterized by low bone density. In the U.S., 10.3% of U.S. adults ≥50 years have osteoporosis^(1)^, with the prevalence highest among postmenopausal women (15.4%)^(1)^. To address this major health problem, optimizing bone accrual in childhood could help reduce the risk of osteoporosis in later life^(2)^.

Our understanding of the polygenic nature of bone health traits, including osteoporosis, has grown extensively during the GWAS era^(3–6)^. While most discoveries have occurred in adults^(3–6)^, we and others have shown that genetic signals for bone health traits operate in childhood^(7–14)^, including polygenic risk scores comprised of GWAS-implicated variants^(15)^. Expanding beyond scores calculated using GWAS-implicated variants, Forgetta et al. applied machine learning to UK Biobank data (adults of European ancestry) to develop a polygenic risk score informed by genetic variants associated with heel quantitative speed of sound data, a measure related to bone mineral density^(16)^. This polygenic risk score, *genetic quantitative ultrasound speed of sound* (gSOS), may enhance screening for osteoporosis in adults^(16)^ and has been associated with reduced odds of fracture in adults of European and Asian ancestry^(17)^.

Only one prior study has investigated gSOS in childhood^(18)^; that study found gSOS scores were lower among a high-risk group of children who had experienced multiple fractures as compared to the general UK biobank adult population^(18)^. Extending these insights, we aimed to determine if gSOS was associated with bone mineral density (BMD) and the likelihood of having a fracture in multi-ancestry samples of children. We hypothesized that a higher gSOS score would be associated with higher BMD and reduced likelihood of fracture.

## Materials and Methods

### Data Sources

Data from the Bone Mineral Density in Childhood Study (BMDCS) examined associations between gSOS and areal BMD (aBMD). Associations between gSOS and fracture were tested using data from the Children’s Hospital of Philadelphia (CHOP) and the BMDCS.

### BMDCS Sample

To establish pediatric bone density reference ranges, the BMDCS was initiated in 2002-2003 and enrolled children aged 6-16 years^(19)^. In 2006-2007, the study was extended to enroll children aged 5 years and adults aged 19 years. Participants were followed annually until 2008-2009, and all were enrolled from one of five sites (Los Angeles, Cincinnati, Omaha, Philadelphia, and New York City)^(19)^. Blood or saliva was collected at the final visit, from which DNA was extracted. In 2010, for a genome wide association study, an additional cross-sectional sample of European ancestry children aged 5-18 years were enrolled at Cincinnati and Omaha. The collection of DNA allowed for this secondary analysis gSOS project. Written informed consent was obtained for participants ≥18 years; participants <18 years provided assent along with consent from a parent or guardian. The Institutional Review Boards at each site approved the protocol.

### BMDCS and gSOS

DNA samples were genome-wide genotyped using Illumina Infinium II OMNI Express plus Exome BeadChips (Illumina, San Diego)^(20)^, and imputed using the 1000 Genome Project. gSOS scores were calculated using 21,716 variants^(21)^.

### BMDCS DXA Methods

Dual energy X-ray absorptiometry (DXA) scans of the lumbar spine, proximal femur, radius and total body were obtained using Hologic (Bedford, MA) bone densitometers (QDR4500A, QDR4500W, Delphi A and Apex models). Scans were analyzed at the University of California, San Francisco’s DXA Core Laboratory. Age and sex-specific Z-scores for aBMD of the total body less head (TBLH), spine, total hip, femoral neck, distal one-third radius, and ultra distal radius were calculated based on age-specific values for the entire BMDCS cohort. Spine bone mineral apparent density (BMAD) Z-scores, to approximate volumetric BMD^(22)^, were also generated^(22)^.

### BMDCS Covariates

Pubertal stage was assessed by physicians or nurses, and participants were categorized as pre-pubertal (Tanner I), pubertal (Tanner II-IV) and post-pubertal (Tanner V)^(23)^. BMI Z-scores were calculated using height (m) and weight (kg) data and U.S. growth charts^(24)^. Dietary calcium intake (g/d) was assessed using a semi-quantitative food frequency questionnaire (Block Dietary Data Systems, Berkeley, CA)^(25)^. Time spent in high-impact, weight bearing physical activity was estimated using a modified version of the Slemenda questionnaire^(26,27)^.

Genetic ancestry was estimated using genotypes of low frequency and common variants that were combined with overlapping SNPs within HapMap Phase 3. The Genome-wide Complex Trait Analysis (GCTA) was used to determine the eigenvalues and eigenvectors of the samples^(28)^. The top 10 eigenvectors were used in a k-nearest neighbor‘s algorithm^(29)^, that was trained on multi-ancestry Hapmap Phase 3 samples, to determine genetic ancestry group.

### Statistical Analyses for gSOS and Bone Density

Linear mixed effect models with random intercepts were used to assess the main effect of gSOS on aBMD and spine BMAD Z-scores. The fixed portion of *model 1* included gSOS and genetic ancestry. In *model 2*, BMI Z-score, dietary calcium and physical activity were added to the fixed portion of model. In *model 3*, sex, Tanner stage, and age were added to the fixed effect portion of the model. To test if gSOS associations were modified by covariates, we included interaction terms between these factors and gSOS in the fixed effect portion of the model. Stata version 17.0 (StataCorp, College Station, TX) was used to perform the modeling.

### CHOP Fracture Sample

Genetic data from the Center for Applied Genomics (CAG) at the Children’s Hospital of Philadelphia were combined with fracture and sex data captured in the electronic health record (EHR). CAG was established in 2006, and enrolled children from the greater Philadelaphia region. For this analysis biologically unrelated individuals were included. The genotype data were imputed using the 1000 Genome Project and gSOS scores were caluclated^15^. This study was approved by the Institutional Review Board of the Children’s Hospital of Philadelphia. Parental informed consent was given for each study participant for both the blood collection and subsequent genotyping.

### CHOP EHR Fracture Data

Cumulative fracture data were extracted from EHR in 2022. Identification of fractures at the 18 most common fracture sites^(30–32)^ was based on ICD9/10 codes (Supplementary Table 1)^(33,34)^.

Participants were categorized as having no history of fracture (referent group) or ≥1 fracture(s). To assess for a dose-response association, participants were categorized as having no history of fracture (referent group), 1 fracture, 2 fractures, or ≥3 fractures. A third variable captured fracture location [no history of fracture (referent group), upper limb fracture, lower limb fracture, or upper & lower limb fracture].

### BMDCS Fracture Data

At enrollment, BMDCS participants reported the number of previous fractures. At each annual visit, they reported fractures that occurred in the prior year^(35)^. Binary and dose-response fracture outcomes as described were generated. At annual visits, a reported the fracture occurrence was categorized as a low (e.g., fall from low height), moderate (e.g., impact during sport) or high energy fracture event (e.g., fall from a high height)^(35)^ allowing particpants to be categorized as having no fracture history (referent), low energy fracture, moderate energy fracture or high energy fracture.

### gSOS and Fracture Statistical Analyses

Logistic regression was used to assess associations between gSOS and the binary fracture outcome. Multinomial logistic regression was used to assess if gSOS was associated with fracture outcomes with four categories. All logistic regression models included sex (fracture rates are higher among male versus female children^(36)^) and genetic ancestry as covariates (fracture rates are lower in children of African versus European ancestry^(35,37–39)^, and gSOS was developed using European ancestry data^(16)^). We also assessed if gSOS associations differed by sex and genetic ancestry by including interactions terms in the models. For the BMDCS fracture sample, models were additionally adjusted for age at last assessment, ultra-distal radius aBMD Z-score and nutrition factors to assess if this impacted any gSOS associations. Finally, BMDCS participants with no prior reported fractures were included in a prospective recurrent fracture event analysis to assess if gSOS was associated with fracture risk. This was done by fitting a Cox model with robust standard errors and accounting for the within-subject correlation, adjusting for the same covariates. Stata version 17.0 (StataCorp, College Station, TX) was used to fit the logistic and Cox models.

## Results

### gSOS and Bone Density

BMDCS sample characteristics are provided (Table 1). Up to 7 annual repeated measures from 1,372 participants were analyzed (7,704 observations). At visit 1, the average age was 11 years, 48% were female, 66% were of European ancestry, and 46% were in Tanner stage I. gSOS was associated with higher aBMD Z-scores at multiple skeletal sites (Figure 1A). Associations were consistent across all models that were incrementally adjusted for covariates (e.g., in *model 3*, a 1 SD increase in gSOS was associated with a 0.17 higher TBLH aBMD Z-score (95% CI: 0.13, 0.21)). The strengths of associations were similar in magnitude across skeletal sites, except for the distal one-third radius where the magnitude was lower (*model 3*, beta=0.08; 95% CI: 0.04, 0.12). gSOS also associated with a marker of volumetric BMD; in *model 3* a 1 SD increase in gSOS was associated with a 0.19 higher spine BMAD Z-score (95% CI: 0.14, 0.24).

**Figure 1.**
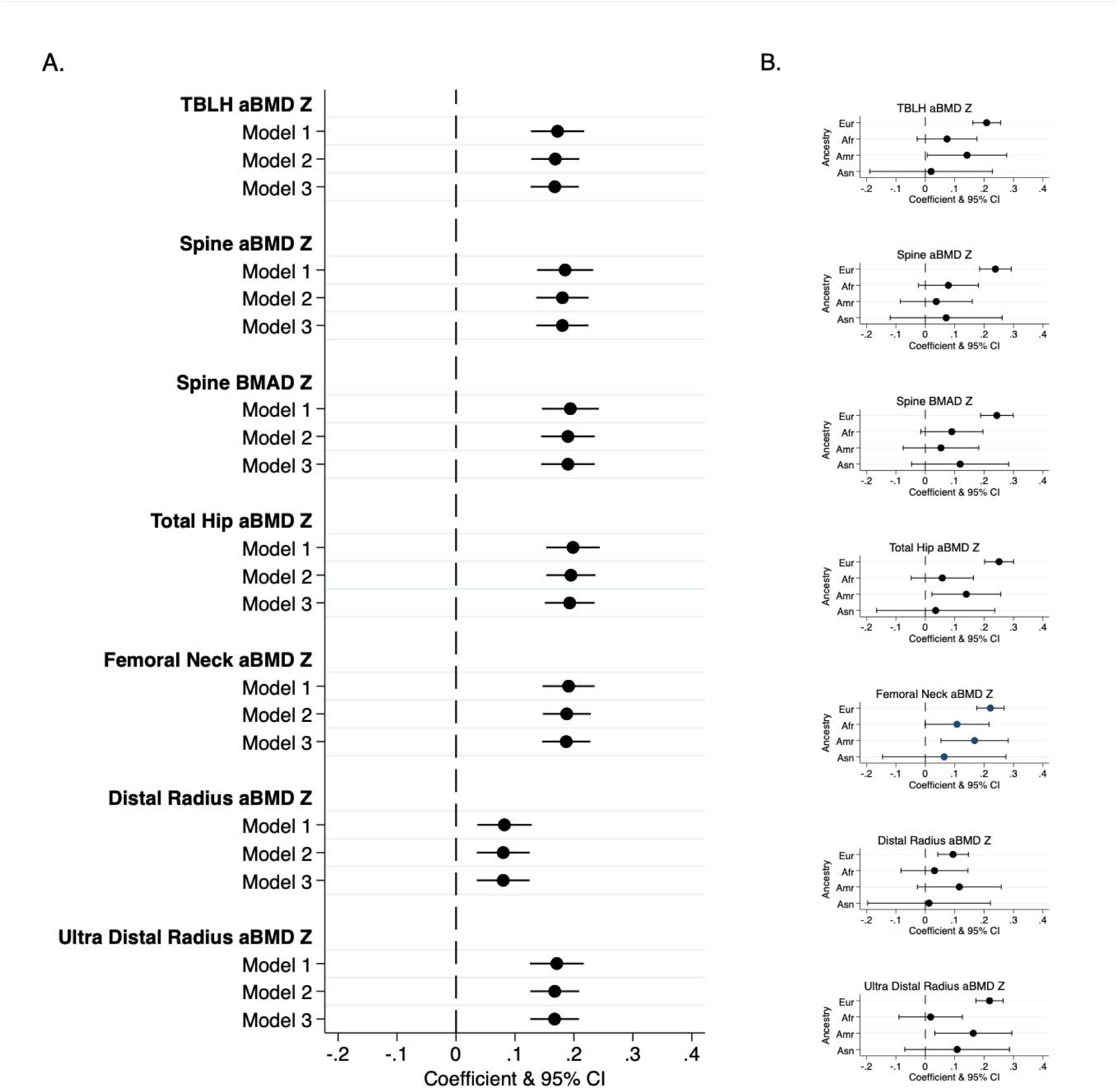
gSOS associations with bone density outcomes for all BMDCS participants (panel A) and by genetic ancestry (panel B). The size and characteristics of the sample used to generate these results are presented in Table 1.

**Table 1.** Descriptive Characteristics of the BMDCS Sample for the aBMD and Spine BMAD Analyses.

|  | Visit 1<br>(N=1,732) | Visit 2<br>(N=1,244) | Visit 3<br>(N=1,233) | Visit 4<br>(N=900) | Visit 5<br>(N=896) | Visit 6<br>(N=879) | Visit 7<br>(N=820) |
| --- | --- | --- | --- | --- | --- | --- | --- |
| Age, y, mean (SD) | 11.4 (4.4) | 12.2 (4.6) | 13.2 (4.5) | 13.7 (3.1) | 14.8 (3.1) | 15.7 (3.1) | 16.6 (3.0) |
| Female, N (%) | 899 (51.9%) | 653 (52.5%) | 647 (52.5%) | 478 (53.1%) | 474 (52.9%) | 463 (52.7%) | 430 (52.4%) |
| European Ancestry, N (%) | 1150 (66.4%) | 669 (53.8%) | 664 (53.9%) | 468 (52.0%) | 465 (51.9%) | 456 (51.9%) | 431 (52.6%) |
| African Ancestry, N (%) | 332 (19.2%) | 330 (26.5%) | 327 (26.5%) | 249 (27.7%) | 248 (27.7%) | 244 (27.8%) | 222 (27.1%) |
| American/Hispanic Ancestry, N (%) | 180 (10.4%) | 177 (14.2%) | 174 (14.1%) | 129 (14.3%) | 129 (14.4%) | 125 (14.2%) | 117 (14.3%) |
| Asian Ancestry, N (%) | 70 (4.0%) | 68 (5.5%) | 68 (5.5%) | 54 (6.0%) | 54 (6.0%) | 54 (6.1%) | 50 (6.1%) |
| Tanner I, N (%) | 796 (46.0%) | 535 (43.0%) | 446 (36.2%) | 194 (21.6%) | 131 (14.6%) | 90 (10.2%) | 145 (17.7%) |
| Tanner II-IV, N (%) | 457 (26.4%) | 285 (22.9%) | 279 (22.6%) | 284 (31.6%) | 269 (30.0%) | 242 (27.5%) | 122 (14.9%) |
| Tanner V, N (%) | 479 (27.7%) | 424 (34.1%) | 508 (41.2%) | 422 (46.9%) | 495 (55.2%) | 547 (62.2%) | 553 (67.4%) |
| Physical Activity, hours per day, mean (SD) | 0.7 (0.7) | 0.6 (0.7) | 0.9 (0.8) | 0.9 (0.8) | 0.9 (0.8) | 0.9 (0.8) | 0.8 (0.8) |
| Dietary Calcium, mg per day, mean (SD) | 905 (514) | 862 (500) | 854 (496) | 843 (534) | 846 (570) | 809 (530) | 773 (476) |
| BMI, Z-score, mean (SD) | 0.3 (0.8) | 0.3 (0.9) | 0.3 (0.9) | 0.3 (0.9) | 0.3 (0.9) | 0.3 (0.9) | 0.3 (0.9) |

We detected interactions between gSOS and genetic ancestry for four aBMD outcomes and spine BMAD (Supplemental Figure 1). The positive gSOS associations with aBMD were most consistent among children of European ancestry and less consistent or null among children of African, Asian, and American ancestry (Figure 1B).

There was no evidence of interactions between gSOS and sex or nutrition factors. However, gSOS association with spine BMAD was incrementally stronger with increasing chronological age (Supplemental Figure 1). Further, the gSOS associations with TBLH aBMD and spine BMAD were partially stronger among post-pubertal versus pre-pubertal children (Supplemental Figure 1).

### gSOS and Likelihood of Fracture

Characteristics of the CHOP cumulative fracture sample are provided (Table 2 and Supplemental Table 2). A 1 SD increase in gSOS was associated with 12% reduced odds of fracture (95% CI: 0.82, 0.95) (Table 3). There was no evidence of a dose-response (Table 3). Regarding fracture site, gSOS was associated with upper limb fracture, but not lower limb fracture (Table 3). Statistical interactions between gSOS and sex or genetic ancestry were not detected (Supplementary Table 3).

**Table 2.** Descriptive Statistics for the CHOP-EHR and BMDCS Samples for Cumulative Fracture Analyses.

|  | CHOP-EHR Fracture Sample<br>(N=10,301) | BMDCS Fracture Sample<br>(N=1,261) |
| --- | --- | --- |
| Female, N (%) | 4,994 (48.5) | 659 (52.3) |
| African Ancestry, N (%) | 5,342 (51.9) | 332 (26.3) |
| American/Hispanic Ancestry, N (%) | 545 (5.3) | 179 (14.2) |
| Asian Ancestry, N (%) | 210 (2.0) | 70 (5.6) |
| European Ancestry, N (%) | 4,204 (40.8) | 680 (53.9) |
| No Fracture, N (%) | 9,242 (89.7) | 950 (75.3) |
| 1 Fracture, N (%) | 581 (5.6) | 244 (19.3) |
| 2 Fractures, N (%) | 344 (3.3) | 52 (4.1) |
| 3 or more Fractures, N (%) | 134 (1.3) | 15 (1.2) |
| No Fracture, N (%) | 9,242 (89.7) | 950 (75.3) |
| Upper Limb Fracture, N (%) | 609 (5.9) | - |
| Lower Limb Fracture, N (%) | 357 (3.5) | - |
| Upper & Lower Limb Fracture, N (%) | 93 (0.9) | - |
| *Unknown Limb, N (%)* | - | *311 (24.7)* |
| No Fracture, N (%) | 9,242 (89.7) | 950 (75.3) |
| Low Energy Fracture, N (%) | - | 29 (2.3) |
| Moderate Energy Fracture, N (%) | - | 108 (8.6) |
| High Energy Fracture, N (%) | - | 25 (2.0) |
| *Unknown Energy, N (%)* | *1,059 (10.3)* | *149 (11.8)* |

**Table 3.**
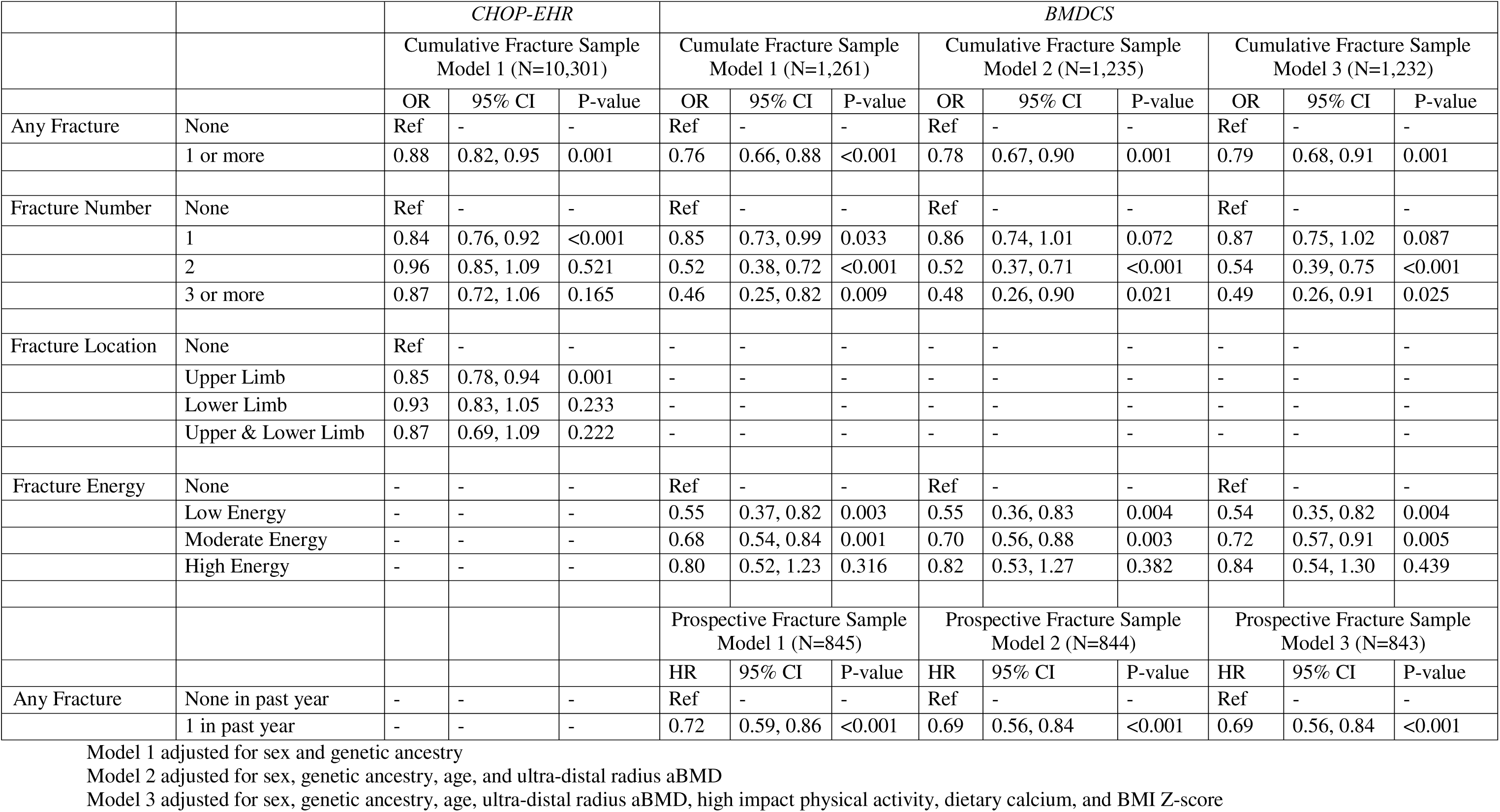
gSOS and Fracture Associations in CHOP and BMDCS Samples.

Characteristics of the BMDCS cumulative fracture sample (Table 2) and the prospective fracture sample are provided (Supplementary Table 3). In the BMDCS cumulative fracture sample, a 1 SD increase in gSOS was associated with 26% reduced odds of fracture (95% CI: 0.66, 0.88) (Table 3), and a dose-response relationship was detected: 15% (95% CI: 0.73, 0.99), 48% (95% CI: 0.38, 0.72) and 54% (95% CI: 0.25, 0.82) reduced odds of one, two, and three or more fractures (Table 3). Further, gSOS was most strongly associated with low energy fracture (OR=0.55, 95% CI: 0.37, 0.82) but was not associated high energy fracture (OR=0.80, 95% CI: 0.52, 1.23) (Table 3). These associations remained with the additional adjustment for age, aBMD and nutrition factors (Table 3). Statistical interactions between gSOS and sex or genetic ancestry were not detected (Supplementary Table 4). However, the gSOS association with fracture was null among children of African ancestry and this was borderline statistically different from children of European ancestry (Supplementary Table 4).

Using the prospective BMDCS fracture sample, with no prior history of fracture, a 1 SD increase in gSOS associated with a 28% reduced risk of fracture (95% CI: 0.59, 0.86), adjusting for sex and genetic ancestry (Table 3). This association remained with the additional adjustment for age, aBMD and nutrition factors (Table 3). Statistical interactions between gSOS and sex were not detected (Supplementary Table 3). However, gSOS association with fracture risk was null among children of African ancestry (Supplementary Table 4). Using the Cox model from Supplementary Table 4, we predicted the proportions of children fracture-free over a 7 year period for those with high versus low gSOS scores for each sex and genetic ancestry (Figure 2). For example, ≈85% of male children of European ancestry were predicted fracture-free after 7 years with a gSOS set at +2SD and this decreased to ≈50% with a gSOS score set at −2SD (Figure 2). This figure also highlights the lower fractures rates among female children and children of African ancestry.

**Figure 2.**
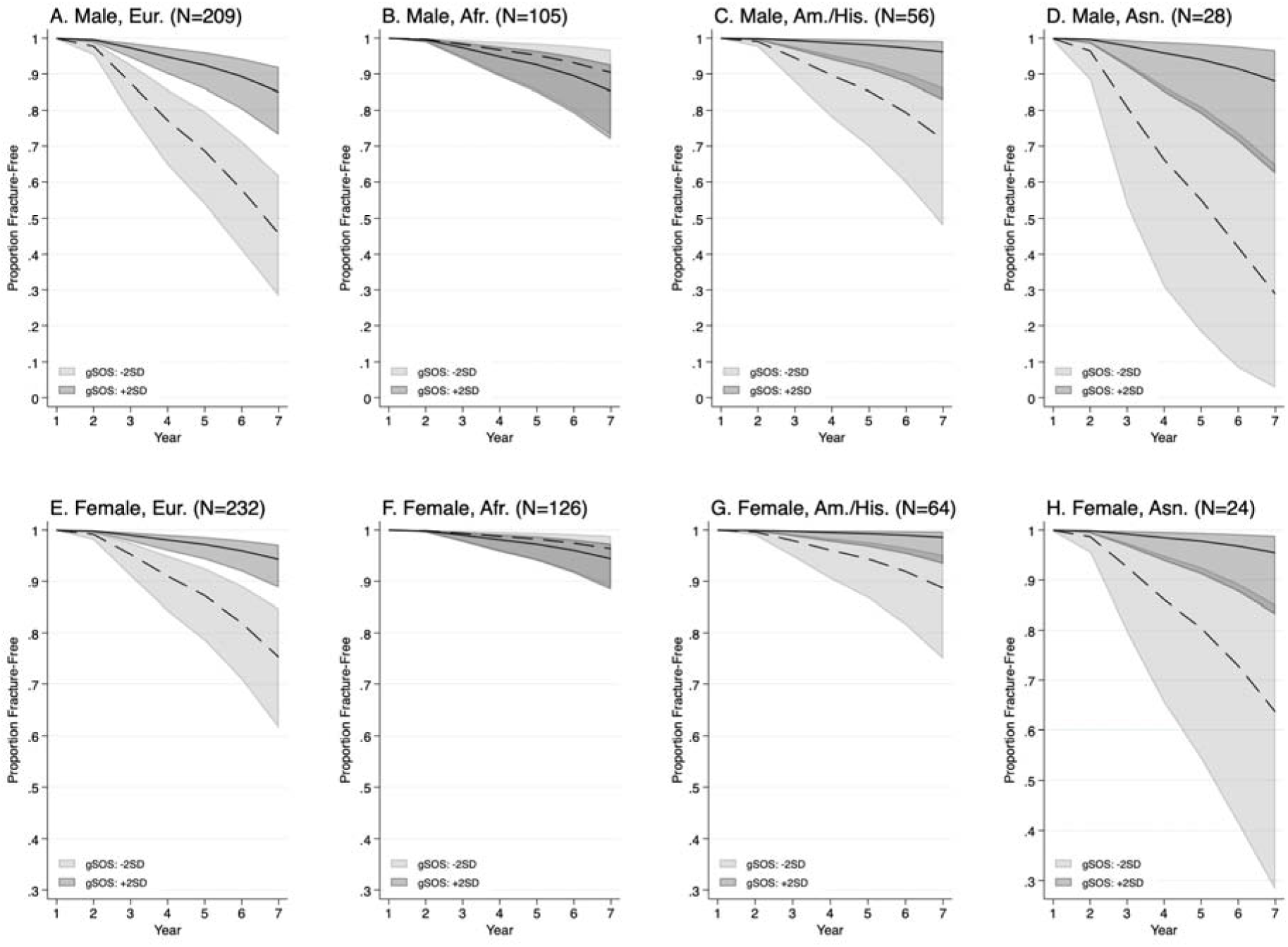
Predicted fracture-free proportions over time for high and low gSOS scores, by sex and ancestry. These predictions are derived from the Cox model in Supplementary Table 4.

## Discussion

The polygenetic risk score gSOS was developed using machine learning applied to genetic variant and heel quantitative ultrasound data from adults of European ancestry^(16)^. This is the first study to demonstrate that gSOS is associated with enhanced bone accrual and fracture reduction in children. We showed: 1) that gSOS associated with higher aBMD across multiple skeletal sites; 2) that gSOS associated with reduced odds of fracture in two independent samples; 3) a dose-response relationship between gSOS and fracture and a particularly strong association with fracture caused by a low energy event were observed; 4) gSOS prospectively associated with reduced risk of fracture; 5) there was no evidence of sex or nutrition-factor differences; and 6) gSOS associations for bone density outcomes were most consistent among children of European ancestry, and while the fracture associations were consistent in all ancestries in cumulative fracture analyses, they were null among children of African ancestry in the prospective BMDCS sample.

We observed consistent positive associations between gSOS and aBMD across multiple skeletal sites, which aligns with estimated bone mineral density data in the gSOS origin study in adults of European ancestry^(16)^. These observations also align with data from young adults who survived childhood acute lymphoblastic leukemia (ancestry unknown)^(40)^, where a 1 SD increase in gSOS associated with a 0.16 increase in lumbar spine aBMD Z-score^(40)^. To the best of our knowledge, no other pediatric study has associated gSOS with aBMD outcomes across multiple skeletal sites. However, children with severe bone disease are reported to have lower gSOS scores compared to adults in the UK Biobank without severe bone disease^(18)^.

The gSOS association with the distal one-third radius aBMD was weaker compared to other skeletal sites, including the ultra-distal radius. The distal one-third radius site is predominantly comprised of cortical bone whereas the ultra-distal site has a higher proportion of trabecular bone^(41)^. gSOS was derived from quantitative ultrasound of the calcaneus which has a high proportion of trabecular bone^(16)^. Therefore, gSOS may operate to protect against fracture through enhanced trabecular bone density and strength. This could be further investigated with more direct measures of cortical and trabecular bone.

In two independent samples, we observed that a 1 SD increase in gSOS was associated with 12% and 26% reduced likelihoods of fracture. These data are similar to adult studies. Across four samples of adults of European ancestry, a 1 SD increase was associated with 26% to 32% reduced odds of fracture^(17)^. That study also reported that among adults of Asian ancestry, a 1 SD increase in gSOS was associated with 11% reduced odds of fracture^(17)^. Additionally, gSOS has been associated with major osteoporotic fracture and hip fracture among adults of European ancestry taking medications known to elevate risk of fracture^(42)^. In contrast, gSOS was not associated with the likelihood of vertebral fracture in a sample of young adults who survived childhood acute lymphoblastic leukemia (ancestry unknown)^(40)^. Limited to the BMDCS, we also report a prospective association between gSOS and a 28% reduced risk of fracture. To our knowledge, there are no pediatric studies to make direct comparisons regarding gSOS and fracture risk in childhood. Further, there are no prior data to draw comparison with our fracture site findings in the CHOP-EHR sample, the dose-response associations in the BMDCS sample, or the fracture energy findings in the BMDCS. Replication is therefore needed.

Osteoporosis and fractures are more common among adult females^(1)^ and childhood fractures are more common among males^(36)^. We found no evidence of sex differences, suggesting gSOS is equally predictive of bone density and fracture in males and females. We also found no evidence of nutritional status differences, indicating gSOS has comparable utility irrespective of a child’s physical activity level, dietary calcium intake, or BMI. Further, this suggests that nutritional recommendations for bone accretion in childhood (e.g., physical activity^(43)^) should be equally effective irrespective of a child’s gSOS determined genetic predisposition to bone fragility. We did, however, observe that gSOS associations may be stronger among older aged and more biologically mature children, which may be a consequence of the adult origins of gSOS. Thus, the machine learning model used to identify genetic variants comprising gSOS should continue to be trained using younger, less biologically mature children.

We found associations between gSOS and aBMD outcomes to be more consistent in children of European ancestry. While not statistically significant in the cumulative CHOP and BMDCS fracture samples, in the prospective BMDCS sample, ancestry differences were detected such that gSOS was not associated with fracture risk in children of African ancestry. This may be partly explained by the low fracture rate among children of African ancestry. Also, there is likely less power to detect ancestry differences with gSOS when using categorical fracture outcomes (as compared to continuous bone density outcomes). However, given that UK Biobank data of adults from European ancestry was used to generate gSOS^(16)^, more consistent associations among children of European ancestry is not unexpected. There are few prior multi-ancestry studies to draw comparisons; however, the gSOS association with osteoporotic fracture was lower among adults of Asian ancestry (11%) compared to adults of European ancestry (26-32%)^(17)^. Additional efforts are warranted to train the underlying machine learning model for gSOS with more diverse data and to test for fracture associations in a larger sample of children of African ancestry.

Our study has limitations. Our genetic findings can only be generalized to ancestries included in the sample; future studies with larger sample sizes across ancestry groups are needed. We do not have data on trabecular and cortical bone density, or structural bone properties. This may explain why the gSOS associations with fracture outcomes in the BMDCS did not attenuate more to the null when adjusting for aBMD. We did not compare and contrast other bone health related polygeneic risk scores; this should be done in the future. In the CHOP-EHR fracture sample, additional fractures experienced by participants may have been recorded at another hospital and in the BMDCS the accuracy of the fracture data was contingent on recall. Additionally, BMDCS participants were ineligible if they had extensive fractures history at enrollment. Further, in the CHOP-EHR sample documentation of ICD9/10 codes were not standardized and cause of fracture was unknown. We also did not consider medications or comorbidities when performing the gSOS associations with fracture outcomes in the CHOP-EHR sample, so the association could be even stronger as this limitation would have biased the gSOS associations towards the null.

In conclusion, a higher gSOS score was associated with higher aBMD at multiple skeletal sites in childhood. In two independent samples, a higher gSOS was protective against fracture and in one sample a dose-response relationship was present, with a particularly strong association with low energy impact fractures, and a prospective reduced risk of fracture was detected. Having a higher gSOS may enhance bone accretion in early life and protect against fracture across the lifespan, and these findings provide the foundation for further investigation into the role of genetic factors in preventing bone fragility throughout the lifecycle.

## Data access

Dr. Mitchell had full access to all the data in the study and takes responsibility for the integrity of the data and the accuracy of the data analysis.

## Disclosures

### Data availability

The data used to generate the results can be made avaiaible upon reasonable request to the corresponding author. In addition, the phenotype data from the BMDCS can be acquired through the NICHD’s Data and Specimen Hub.

### Ethics approval statement

Ethical approval for this study was obtained from The Children’s Hospital of Philadelphia Committee for the Protection of Human Subjects protocol numbers 02-002724, 06-004886, and 10-007544.

---

**Supplemental Figure 1.**
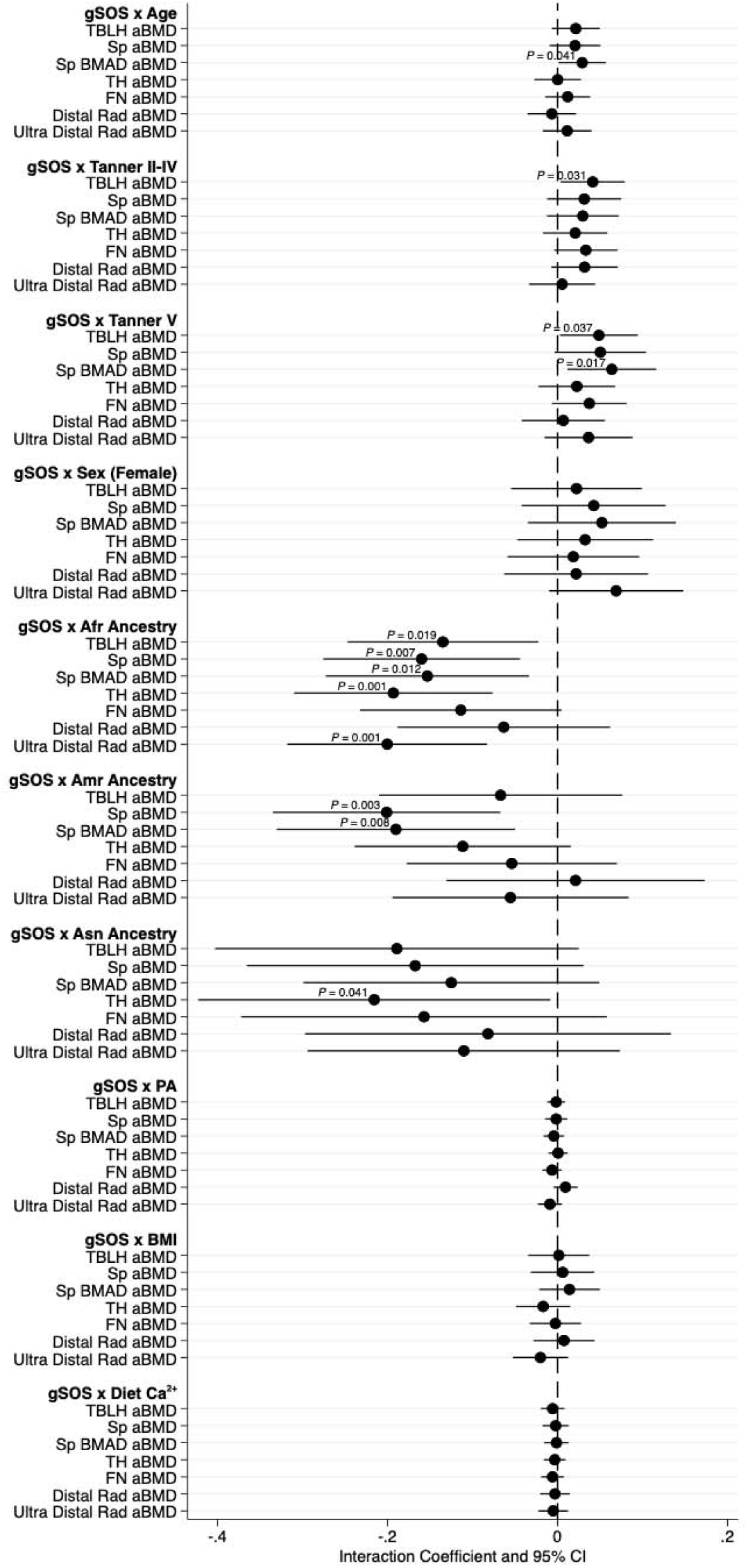
Statistical interactions between gSOS and demographic and lifestyle factors with respect to bone density outcomes in the BMDCS.

**Supplementary Table 1.** The ICD9 and ICD10 Codes for Fracture Identification in the CHOP EHR.

| **Bones** | **ICD9 (fragility)** | **ICD10 (non-fragility)** | **ICD10 (fragility)** |
| --- | --- | --- | --- |
| Ankle |  | S82.5, S82.6 | M84.471, M84.472, M84.473 |
| Clavicle |  | S42.0 |  |
| Fibula |  | S82.4 | [subset of M84.46] |
| Humerus | 733.11 | S42.2, S42.3, S42.4 | M84.42 |
| Metacarpal |  | S62.1, S62.2, S62.3 |  |
| Metatarsal |  | S92.0, S92.1, S92.2 | M84.474, M84.475, M84.476 |
| Neck of Femur | 733.14 | S72.0 |  |
| Femur | 733.15 | S72.1, S72.2, S72.3, S72.4, S72.8, S72.9 | M84.451, M84.452, M83.453 |
| Patella |  | S82.0 |  |
| Pelvis |  | S32.1, S32.2, S32.3, S32.4, S32.5, S32.6, S32.8 | M84.454 |
| Radius |  | S52.1, S52.3, S52.5 | [subset of M84.43] |
| Radius & ulna | 733.12 | S52.9 | M84.43 |
| Rib |  | S22.2, S22.3, S22.4, S22.5, S12.8 |  |
| Scaphoid |  | S62.0 |  |
| Tarsal |  | S92.3 | [subset of M84.474, M84.475, M84.476] |
| Tibia |  | S82.1, S82.2, S82.3 | [subset of M84.46] |
| Tibia & fibula | 733.16 |  | M84.46 |
| Ulna |  | S52.0, S52.2, S52.6 | [subset of M84.43] |

**Supplementary Table 2.** Fracture site frequency data for the cumulative CHOP-EHR Sample.

| Site | Location | N |
| --- | --- | --- |
| Radius/Ulna | Upper | 377 |
| Humerus | Upper | 202 |
| Metacarpal | Upper | 187 |
| Tibia/Fibula | Lower | 185 |
| Tibia | Lower | 182 |
| Tarsal | Lower | 181 |
| Metatarsal | Lower | 179 |
| Scaphoid | Upper | 97 |
| Femur | Lower | 81 |
| Femoral Neck | Lower | 22 |
| Pelvis | Lower | 21 |
| Patella | Lower | 18 |
| Ribs | Upper | 17 |

**Supplementary Table 3.** Descriptive Statistics for the BMDCS Prospective Fracture Sample.

|  | Visit 1<br>(N=844) | Visit 2<br>(N=813) | Visit 3<br>(N=789) | Visit 4<br>(N=685) | Visit 5<br>(N=662) | Visit 6<br>(N=632) | Visit 7<br>(N=589) |
| --- | --- | --- | --- | --- | --- | --- | --- |
| Age, y, mean (SD) | 10.5 (3.6) | 11.5 (3.6) | 12.5 (3.6) | 13.4 (3.0) | 14.4 (3.0) | 15.3 (3.0) | 16.3 (3.0) |
| Female, N (%) | 446 (52.8) | 428 (52.6) | 414 (52.5) | 361 (52.7) | 347 (52.4) | 330 (52.2) | 303 (51.4) |
| European Ancestry, N (%) | 441 (52.3) | 421 (51.8) | 408 (51.7) | 331 (48.3) | 318 (48.0) | 301 (47.6) | 283 (48.0) |
| African Ancestry, N (%) | 231 (27.4) | 224 (27.6) | 215 (27.2) | 201 (29.3) | 196 (29.6) | 185 (29.3) | 169 (28.7) |
| American/Hispanic Ancestry, N (%) | 120 (14.2) | 118 (14.5) | 116 (14.7) | 105 (15.3) | 100 (15.1) | 98 (15.5) | 92 (15.6) |
| Asian Ancestry, N (%) | 52 (6.2) | 50 (6.2) | 50 (6.3) | 48 (7.0) | 48 (7.3) | 48 (7.6) | 45 (7.6) |
| Physical Activity, hours per day, mean (SD) | 0.6 (0.7) | 0.5 (0.7) | 0.9 (0.8) | 0.9 (0.7) | 0.9 (0.8) | 0.9 (0.8) | 0.8 (0.8) |
| Dietary Calcium, mg per day, mean (SD) | 918 (521) | 859 (487) | 868 (500) | 832 (516) | 854 (563) | 816 (529) | 789 (475) |
| BMI, Z-score, mean (SD) | 0.3 (0.8) | 0.3 (0.9) | 0.3 (0.9) | 0.3 (0.9) | 0.3 (0.9) | 0.3 (0.9) | 0.3 (0.9) |
| Fractures in Past Year, N (%) | 0 | 14 (1.7) | 24 (3.0) | 21 (3.1) | 28 (4.2) | 22 (3.5) | 24 (4.1) |

**Supplementary Table 4.**
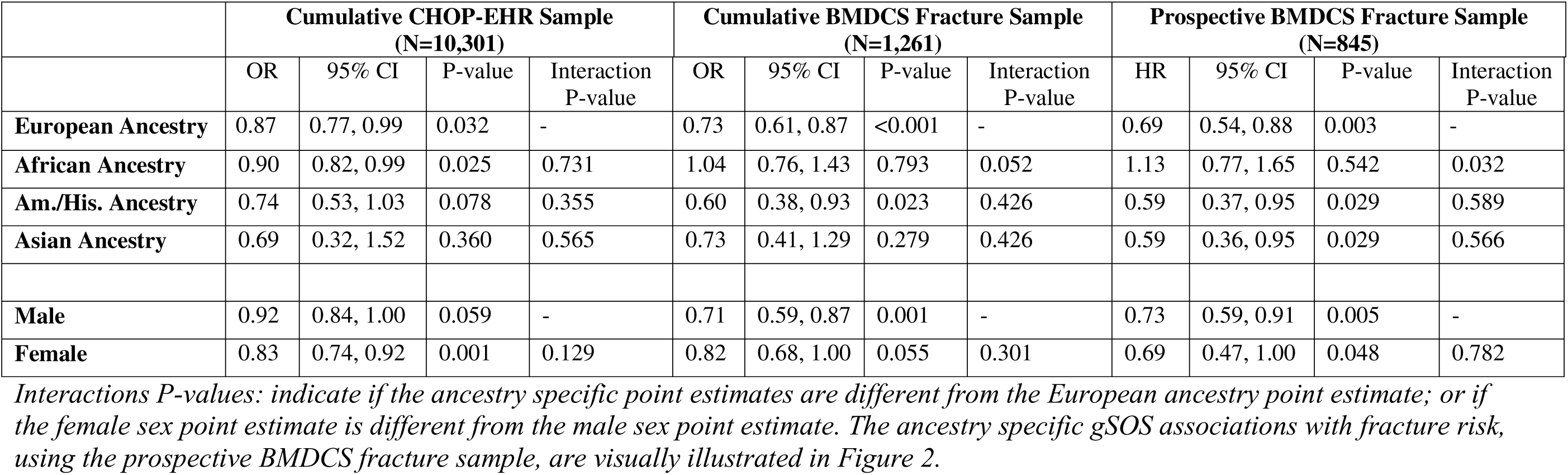
Tests of sex and ancestry differences regarding gSOS associations with fracture.

